# Carbon monoxide exposure in pregnant women in the UK – results from the IPPCO study

**DOI:** 10.1101/2025.01.07.25320119

**Authors:** Elsie Place, Hilary Wareing, Mari Herigstad

## Abstract

Carbon monoxide (CO) is a colourless, odourless gas that poses a threat to life at concentrations of just a few hundred ppm. The developing foetus is particularly vulnerable to CO exposure, and maternal exposure to much lower levels of the gas is associated with adverse outcomes such as low birth weight. This study aimed to quantify CO exposure in pregnant women’s homes and assess whether breath CO levels could be linked to home-based CO exposure and sociodemographic factors.

CO levels were monitored continuously over two weeks in 161 households selected for indicators of lower socio-economic status and proximity to gas appliances, a risk factor for environmental CO exposure. Exhaled breath CO measurements were taken before and after the monitoring period. Of the households monitored, 57.8% had detectable CO levels, with 31.7% experiencing levels above 4ppm and 14.3% above 10ppm. CO exposure varied significantly across households, with both chronic low-level and intermittent high-level exposures observed. Six households included in the study exceeded current World Health Organisation recommended limits of 3.5ppm for ≥24 hours, and three exceeded the limit of 9ppm for ≥8 hours. Higher CO levels in the household were associated with the use of gas for cooking. Higher exhaled CO levels were associated with number of smokers in the household and eligibility for the UK government NHS Healthy Start scheme. Following the monitoring period, exhaled CO levels were only associated with number of smokers in the household, suggesting an intervention effect.

This study indicates that a high proportion of pregnant women are exposed to CO within the home, albeit predominantly within current recommended limits, and that exposure may be linked to socio-economic factors in addition to smoking. This study highlights the need for improved CO monitoring and mitigation strategies, particularly in vulnerable populations, to protect maternal and foetal health.

## INTRODUCTION

Carbon monoxide (CO) is a colourless, odourless, and tasteless gas which binds readily to haemoglobin in the blood, forming carboxyhaemoglobin (COHb) at the expense of oxyhaemoglobin (OHb; (D. Penney et al., 2010). CO also shifts the OHb dissociation curve, leading to potentially lowered oxygen release to tissues and hypoxia (Longo, 1977; Raub & Benignus, 2002; Townsend & Maynard, 2002). Toxic effects beyond hypoxia may also contribute to ongoing morbidity, as symptoms may persist or emerge even after COHb levels normalize (Thom et al., 1995). Acute CO poisoning in pregnant women is linked to complications such as preterm birth and miscarriage, with the outcomes of pregnancy generally being influenced by the severity of maternal poisoning and the stage of foetal development (Smollin & Olson, 2008). However, the developing foetus is uniquely vulnerable to disruptions, and at elevated risk of CO poisoning due to foetal Hb having a higher affinity for CO than adult Hb, a risk that continues into the neonatal period as foetal Hb persists for approximately 6 months after birth. Chronic exposure to subacute levels of CO – particularly in the context of maternal smoking – is also associated with a range of adverse foetal outcomes (Batstra et al., 2003; Dadvand et al., 2011; Dix-Cooper et al., 2012; Lee et al., 2011; Pineles et al., 2014; Ritz et al., 2002; Venditti et al., 2013). Alarmingly, epidemiological studies have revealed negative associations between ambient (environmental) CO levels and birth weight, and linked CO to an increased risk of intrauterine growth restriction (Ritz & Yu, 1999; Salam et al., 2005; Thompson et al., 2011; Trentini et al., 2016), in both cases at average CO parts per million (ppm) concentrations in the single figures. Animal studies have shed further light on CO effects, showing that exposures leading to maternal COHb levels associated with smoking lead to behavioural and brain histochemical abnormalities in offspring (Cagiano et al., 1998; Di Giovanni et al., 1993; Giustino et al., 1999; Trentini et al., 2016), and recently linking low (≤18ppm) CO levels to changes in the developing heart (Matias et al., 2024). Therefore, there is some uncertainty regarding safe CO levels for the unborn child. Reflecting the potential for harm at even single digit concentrations, current safety guidelines state that individuals should not be exposed to an excess of 9 ppm for 8 hours (or more), or 3.5 ppm for 24 hours (or more, WHO, 2021), substantially lower than the levels generally associated with toxic symptoms in adults.

In 2010, the National Institute for Health and Care Excellence (NICE) issued guidelines recommending the use of exhaled CO as an indicator of smoking in pregnant women (NICE, 2010). Consequently, midwives became involved in monitoring CO levels in pregnant women, leading to an observation that elevated breath CO levels are present in a subset of non-smoking pregnant women. This suggested that these women were exposed to alternate sources of CO, potentially including the home and/or urban environment, and that there remains uncertainty regarding the scale of CO exposure among pregnant women and therefore its potential impacts.

To address these issues, a multi-site research project was conducted across five NHS hospital sites providing maternity services. Participants included pregnant women and healthcare professionals involved in their care, selected based on specific inclusion criteria. Study sites were chosen to include those serving deprived populations, as women from such areas might have less access to CO monitoring equipment and may experience higher levels of exposure due to suboptimal housing conditions and appliance malfunctions. The project aimed to quantify CO levels in the homes of pregnant women through a two-week monitoring period and to assess whether breath testing at booking could serve as an effective indicator of home-based CO exposure. Additionally, the study sought to collect household data to identify CO sources and evaluate exposure levels in relation to potential health risks. Demographic data were also correlated with CO exposure levels to provide a more comprehensive analysis. CO was detected in over half of the study households, and in over 5% of households (9/161) levels breached current World Health Organisation (WHO) safety guidelines. Higher CO levels were associated with the use of gas cooking appliances, and breath CO levels were mainly associated with smoking. These findings highlight potential risks to maternal and foetal health posed by subacute CO exposure from sources other than smoking.

## MATERIALS AND METHODS

Participants were recruited from maternity services at the University Hospital Coventry & Warwickshire, Royal United Hospital Bath, Queen Elizabeth Hospital Gateshead, Mid Yorkshire NHS Trust (Pinderfields and Pontefract Hospitals) and Northumbria Hospital. Pregnant women aged 18 years or older, presenting for care at a booking visit or in the early stages of their pregnancy, were eligible for the study. The inclusion criteria were: pregnant; currently living in private rented accommodation or social housing or eligible for healthy start vouchers; living in a property with gas heating/cooker and/or solid fuel or oil and/or living above a takeaway/café/restaurant; able to provide informed consent for the study. Women from all ethnic backgrounds were considered and interpreter services were available for the consent and data collection process to aid inclusivity. Ethical approval was obtained (IRAS ID 301261).

### Protocol

The study included two experimental home visits (outlined below) conducted by the participant’s local Fire and Rescue Service (FRS) and a two-week sampling period.

### Visit 1

A CO alarm (AICO Ei208, AICO, Shropshire) was fitted in the property (if one was not already present) and the ambient CO level was recorded. An expired breath CO test obtained from the participant using Bedfont Toxco Breathalysers (Bedfont Scientific Ltd, Kent, England) according to the manufacturer’s instructions. CO data loggers (EL-USB-CO, Lascar Electronics, Wiltshire, UK), which measure and store over 32k CO readings over a 0 to 1000 ppm range, were placed in each property at the area of the highest perceived risk.

### Sampling period

The CO data logger remained in the property for a minimum of two weeks, recording ambient CO (one reading every 5 minutes).

### Visit 2

Following the two-week sampling period, the dataloggers were collected by the FRS and the anonymised data downloaded. During this visit, the CO level was recorded as measured by the CO alarm, and a further breath test was taken. A questionnaire (collected manually or using a laptop) was completed by the FRS using information provided by the pregnant women and observations made whilst in the home. The questionnaire covered household details including form of housing and residents’ status, as well as the presence of any appliance(s) and fuel type(s) (Kokkarinen et al., 2014). Women were not asked specifically whether they were smokers as part of this study. Table 1 outlines all parameters recorded in this study.

**Table 1.**
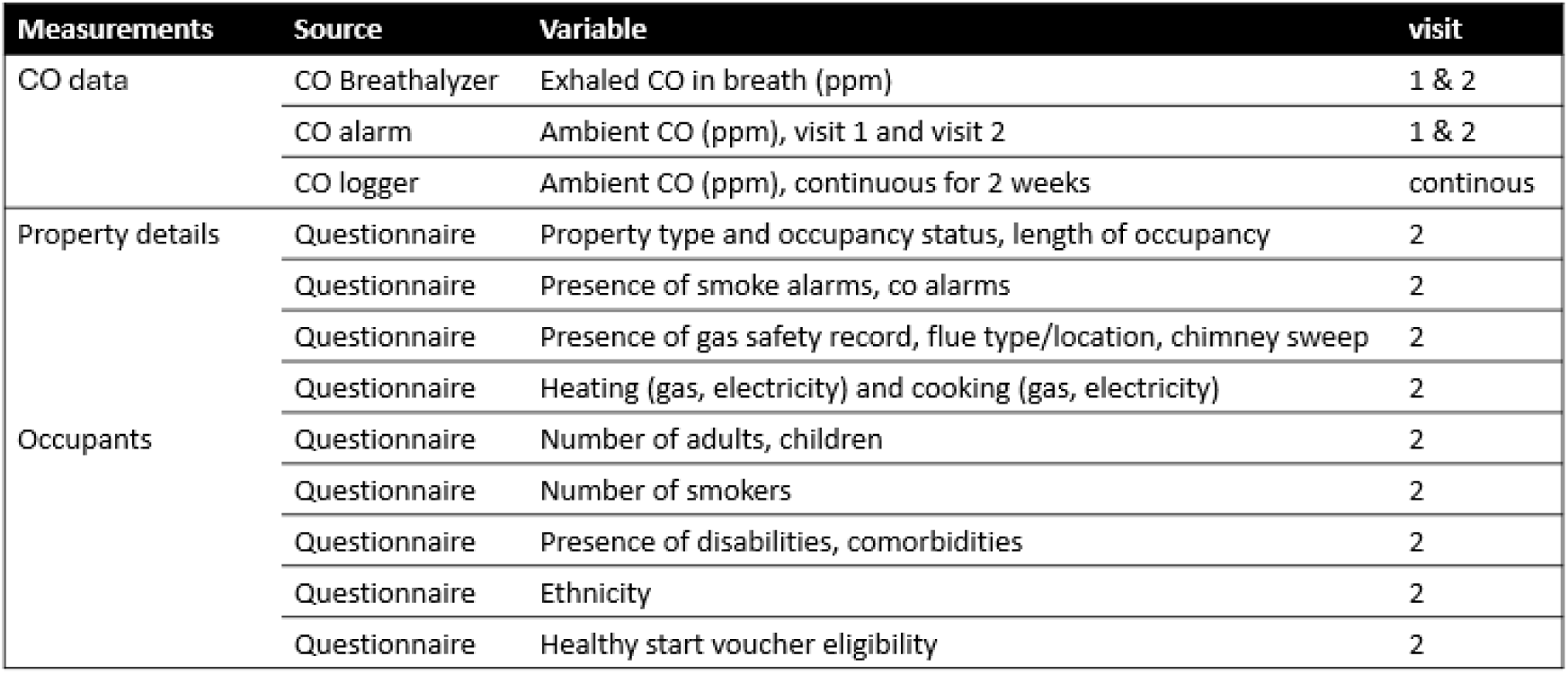
Data collected, sources and variables.

### Data analysis and statistical comparisons

The readings from the data loggers (CO log timeseries data) were downloaded using EasyLogUSB, Lascar’s own software (Lascar Electronics, 2021), and converted to .txt files with a unique participant identification number. All CO log files were then truncated at two weeks, to ensure uniform sampling time. No file was shorter than two weeks. All other data was collated on a spreadsheet (Microsoft Ltd, Redmond, US) using the same identification code.

Timeseries data was analysed using custom-written software (MATLAB, Mathworks Inc, Natick, US) to quantify overall maximum (max) CO exposure levels, overall average (mean) CO exposure levels, and characteristics of any prolonged CO exposures (long exposure, LE, defined as CO above zero at each sampling point for 10 minutes or more), including total duration of LEs (in minutes), max LE duration (in minutes), mean LE duration (in minutes), and max and mean CO levels during LEs.

Timeseries data was also analysed for mean and max CO exposure levels across time of day, to generate a profile of CO exposure fluctuations on an hour-by-hour basis. This was done by averaging the CO levels across each hour (e.g. 01:00-01:55) for each individual household and assigning them to the hour for all days, then averaging across the two-week logging period. This method is a conservative assessment of fluctuations, as it did not take into account any behavioural factors, such as different routines during the week versus weekend. Outputs were visually inspected for any trends and a curve estimation was conducted (IBM SPSS Statistics, Version 27).

CO measurements (exhaled breath data, data from alarms) were compared before and after the sampling period (at first and second visit) using a Student’s t-test (two tailed, paired). A Shapiro-Wilk test of normality was done on all variables, followed by an explanatory correlation analysis (IBM SPSS) to compare CO exposure levels (derived from CO log timeseries data), other CO measurements obtained (breath measurements, alarm values), and all demographic data (bivariate correlation). A subsequent stepwise linear regression analysis was conducted to identify the most significant predictors of the CO level at visit 1 (exhaled breath) from the demographic data (independent variables). The criteria for entry and removal of variables were based on a significance level of 0.05. Variance Inflation Factors for all predictors were accepted if below 2. This was conducted to ascertain whether exhaled CO in the sample could be predicted by any risk factors for exposure.

## RESULTS

Only complete data sets were included in the analysis. CO log data as well as data from visits 1 and 2 were collected in 161 participants (Table 2).

**Table 2:**
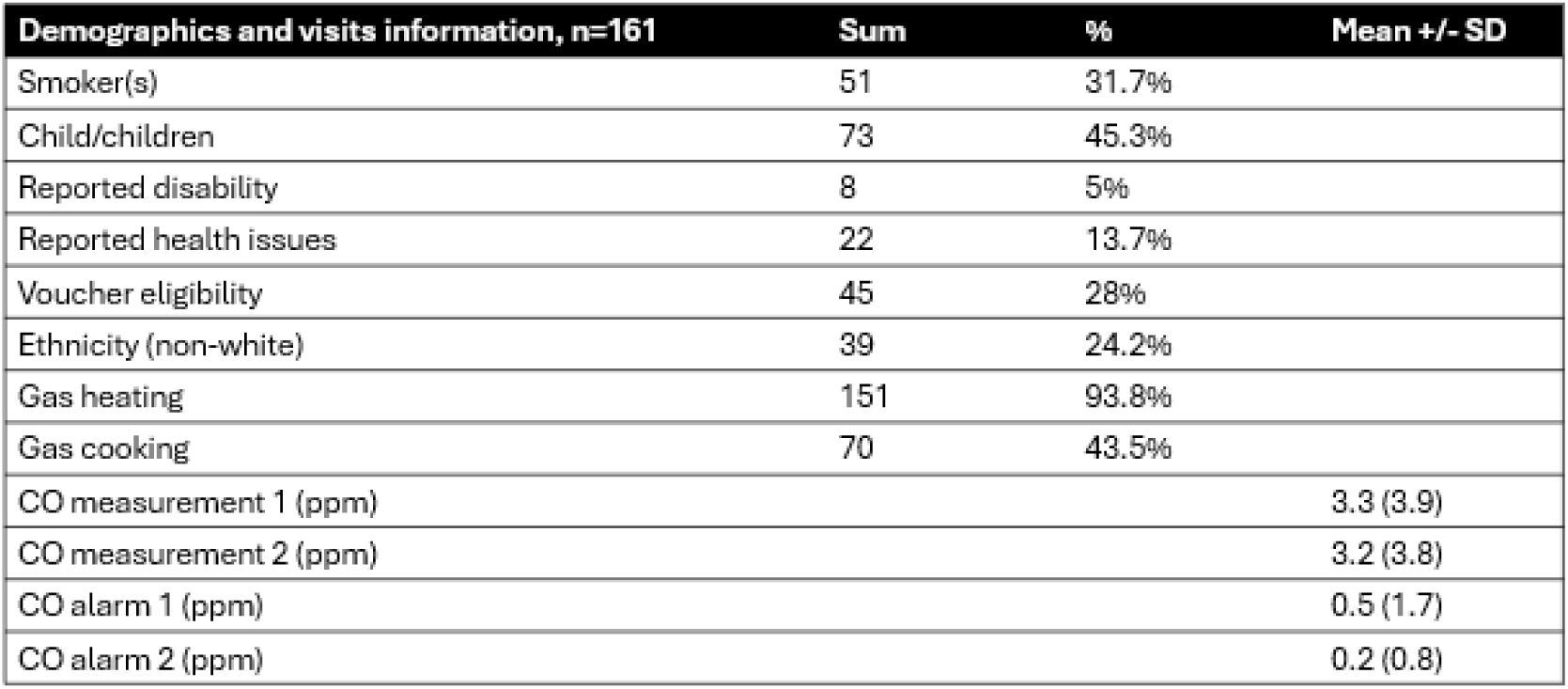
Data from visits 1 and 2 (n=161).

| Demographics and visits information, n=161 | Sum | % | Mean +/- SD |
| --- | --- | --- | --- |
| Smoker(s) | 51 | 31.7% |  |
| Child/children | 73 | 45.3% |  |
| Reported disability | 8 | 5% |  |
| Reported health issues | 22 | 13.7% |  |
| Voucher eligibility | 45 | 28% |  |
| Ethnicity (non-white) | 39 | 24.2% |  |
| Gas heating | 151 | 93.8% |  |
| Gas cooking | 70 | 43.5% |  |
| CO measurement 1 (ppm) |  |  | 3.3 (3.9) |
| CO measurement 2 (ppm) |  |  | 3.2 (3.8) |
| CO alarm 1 (ppm) |  |  | 0.5 (1.7) |
| CO alarm 2 (ppm) |  |  | 0.2 (0.8) |

31.7% of households included at least one smoker, 45.3% included children, 5% included individuals with disabilities, and 13.7% included individuals with reported health issues (not specified). 28% were eligible for healthy start vouchers, and 24.2% were of non-white ethnicity. 93.8% used gas for heating as opposed to non-gas sources, and 43.5% used gas for cooking, as opposed to non-gas sources. There was no difference in CO breath measurements (p=0.94) nor CO alarm measurements (p=0.06) before and after the sampling period.

### CO log timeseries analysis

Representative CO traces highlighting different patterns of exposure are presented in Figure 1. In some households, chronic low-level CO was measured across the monitoring period (Fig 1A). In others, the exposure was more intermittent with either no discernible pattern (Fig 1B) or a clear pattern of exposure (Fig 1C). Levels varied greatly both across and within households, with some experiencing repeated or continued exposures of 10ppm and (considerably) above (Fig 1C).

**Figure 1.**
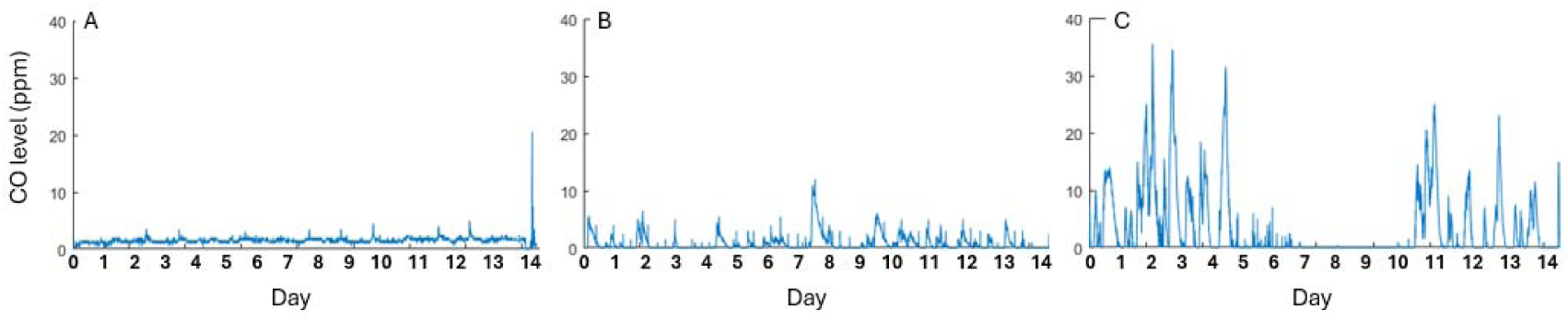
Example CO log timeseries data across 14 day assessment periods, showing (A) chronic, low-level exposure (<3.5ppm), (B), intermittent moderate exposure (<9.0ppm) without a clear pattern and (C) intermittent, high exposure (>9.0ppm) with a clear pattern. Bars at the bottom of the graph indicate mid-point of each day of assessment.

Further assessment of the timeseries showed that CO levels typically were lower during the day than the night (Fig 2A), and that there was a spike in CO levels around 17:00 – 18:00 (Fig 2B). Curve estimation showed that the best fit for the data was a polynomial cubic model.

**Figure 2.**
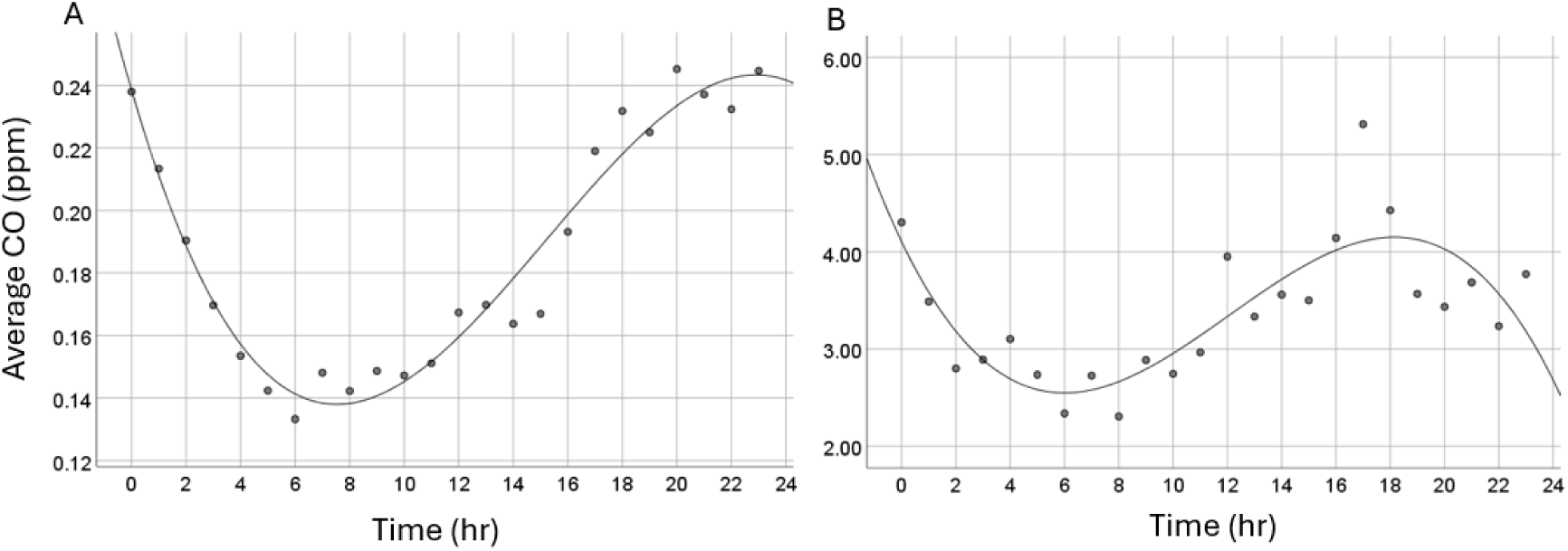
Mean (A) and max (B) CO values over the 24-hour period. Data are averages for each hour across two weeks, pooled across all samples. Data is fitted with a polynomial trendline for mean (R2 cubic = 0.95, p<0.001) and max (R2 cubic = 0.81, p<0.001).

CO log timeseries analysis showed that 57.8% of households (93 households) had at least one CO reading above zero at some point during the sampling period. 31.7% of the households experienced CO levels above 4ppm at least once (51 households), 16.7% experienced CO levels above 8ppm and 14.3% experienced CO levels above 10ppm at least once (27 households and 23 households, respectively). On average, for those exposed, time spent >4ppm was 9hrs (544.2+/-1543 min), >8ppm was 4 hrs (237.4+/-718.8 min) and >10ppm was 3.3 hrs (198.0+/-606.3 min, not necessarily consecutive exposure) over the two-week period. 47.8% of all households had exposures (CO above zero) that lasted 10 minutes or more (LEs). Three households exceeded the WHO exposure guideline of 9ppm over 8 hours and six households exceeded the WHO exposure guideline of 3.5ppm over 24 hours during the course of the study.

In households where CO was detected, the average exposure across the entire two weeks sampling period was 0.3+/-0.7ppm and the average maximum was 10.4+/-33.5ppm. Households with LEs were on average exposed 23.5 times, with a mean duration of such LEs being 533+/-2345mins and a median duration of 55mins (interquartile range (IQR) of 100mins: 33mins (Q1) -133mins (Q3)). Average CO level during LEs was 1.5+/-2.3ppm.

Correlations (Spearman’s rho) for all variables are presented in Figure 3D, and CO-related correlations significant at the p<0.01 level are described in text. We observed that exhaled CO levels at visit 1 (breath1) correlated strongly with exhaled CO levels at visit 2 (breath2; r=.54, p<0.001), with number of smokers in the property (# smokers; r=0.41, p<0.001) and with voucher eligibility (not eligible vs eligible; r=.25, p=0.002). Thus, breath1 levels were higher in those with higher breath2 levels, higher number of smokers in the household and who were eligible for healthy start vouchers. Exhaled CO levels (breath2) correlated with number of smokers (r=.27, p=0.001). Thus, breath2 levels were higher in those with higher number of smokers in the household.

**Figure 3.**
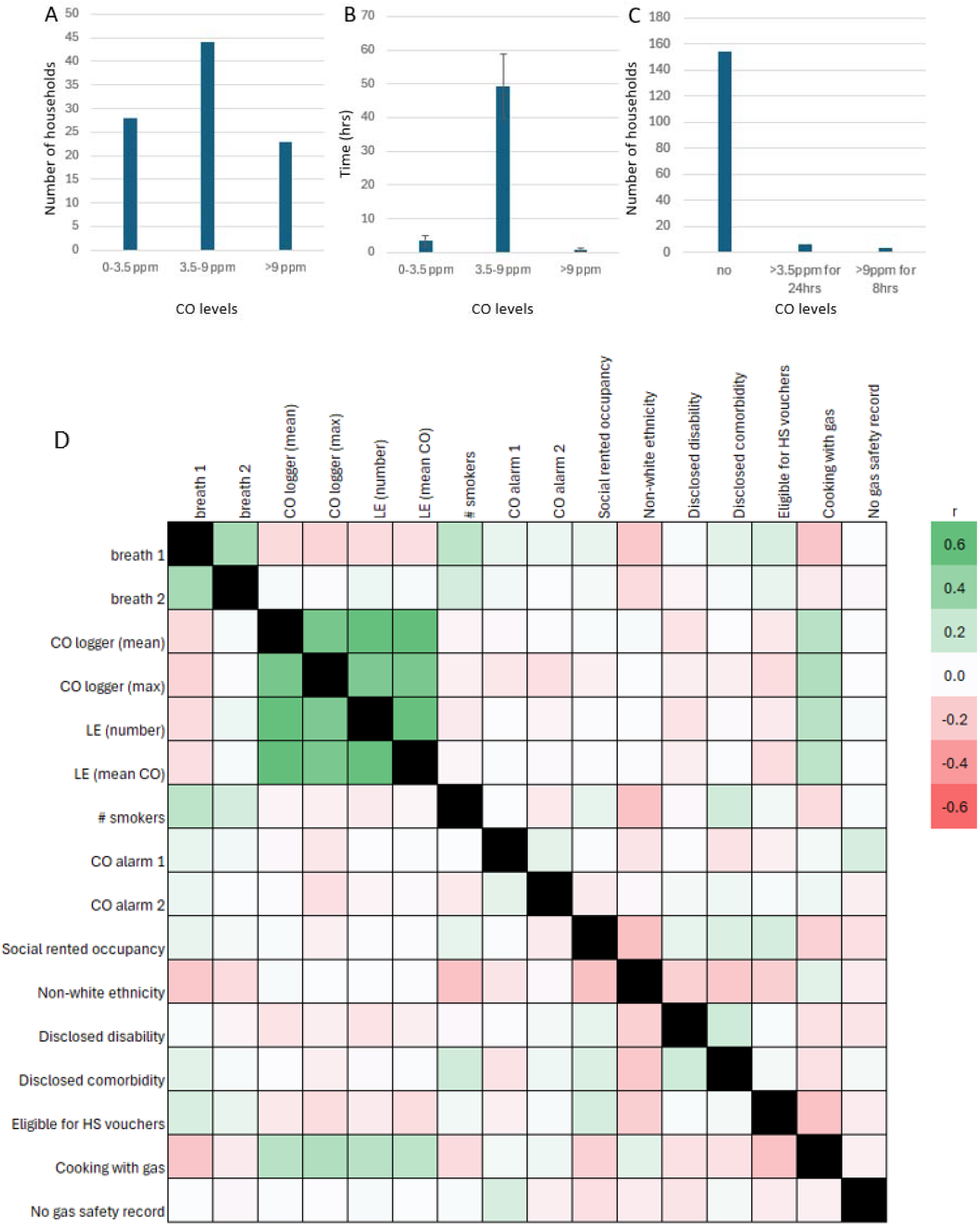
CO log data. (A) Number of households with CO log data showing CO exposures at 0-3.5ppm, 3-9ppm and >9ppm at least once during the study period; (B) average time (not necessarily consecutive) at CO exposures of 0-3.5ppm, 3-9ppm and >9ppm, average +/-SE; (C) number of households adhering to or exceeding WHO guidelines; (D) Exploratory analysis demonstrating correlations of changes in the measured variables across the sample (full correlation matrix). Stronger correlations are highlighted in red (negative) and green (positive correlations). r = Spearman’s rho. HS = Healthy Start.

Mean CO levels collected by the data loggers (CO logger (mean)) correlated with maximal CO levels (CO logger (max); r=0.81, p<0.001), number of LEs (LE (number); r=0.91, p<0.001) and mean CO during LEs (LE (mean CO); r=0.92, p<0.001). CO logger max correlated with LE number (r=0.77, p<0.001) and LE mean CO (r=0.79, p<0.001), and LE number correlated with LE mean CO (r=0.91, p<0.001). Thus, CO logger parameters correlated positively with each other.

Heating method yielded no data as > 90% of households used gas for heating, meaning there was too little variation in the sample for analysis. Cooking with gas correlated with all CO log data: mean CO levels (r=0.43, p<0.001), max CO levels (r=0.47, p<0.001), number of LEs (r=0.43, p<0.001) and mean CO levels during LEs (r=0.40, p<0.001). Finally, having a gas safety record correlated with alarm level on visit 1 (alarm1; r=0.25, p<0.006).

We conducted a linear, stepwise regression analysis to determine predictors of exhaled CO levels. The most important predictors of exhaled CO levels at visit 1 were number of smokers in the household and voucher eligibility. As CO log data correlated poorly with exhaled CO data (Figure 3), log data were not entered into the model. The final model explained a significant proportion of the variance in exhaled CO levels (R^2^=0.31, adjusted R^2^=0.25, F(2,89)=19.09, p<0.001). Coefficients for the predictors are presented in Table 3. Repeating the prediction analysis for breath2 showed that only smoking was a predictor for exhaled CO at visit 2 (breath2).

**Table 3:** Stepwise regression analysis.

| Predictor | Coefficient (B) | Standard Error (SE) | t-value | p-value |
| --- | --- | --- | --- | --- |
| Constant | -0.755 | 1.212 | -0.623 | 0.535 |
| # Smokers | 2.988 | 0.574 | 5.208 | 0.000 |
| Voucher eligibility | 2.381 | 0.908 | 2.622 | 0.010 |

There was a statistically significant difference between exhaled CO levels (breath1) in households with no, one and more than one smokers, as determined by one-way ANOVA (F(2,158) = 28.008, p<0.001; Figure 4A). There was a statistically significant difference between exhaled CO levels in households that were eligible and not eligible for Healthy Start vouchers, as determined by Student’s T-test (p=0.002; Figure 4B).

**Figure 4.**
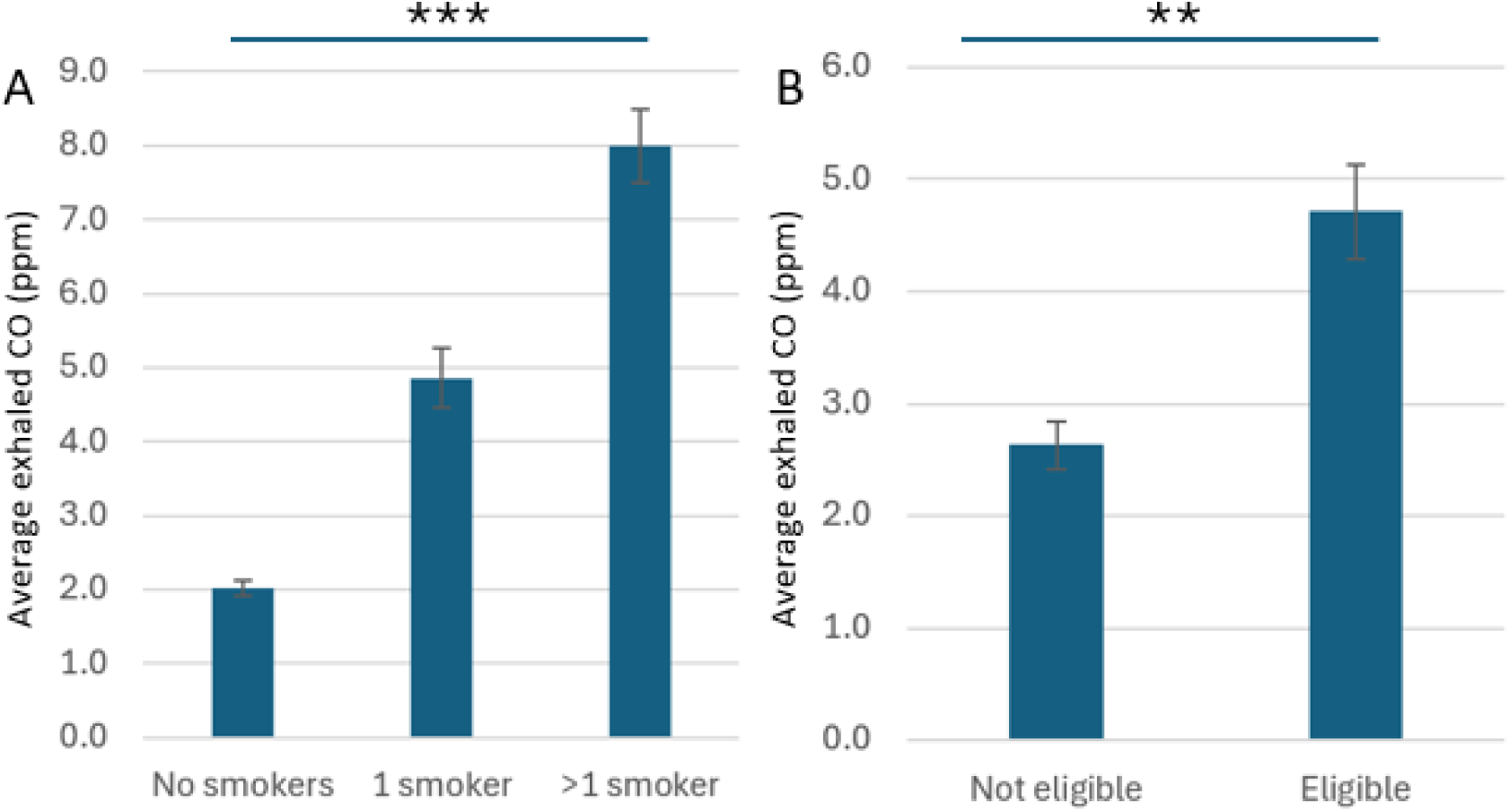
Exhaled CO (first visit) in pregnant women living in (A) households with no, one or more than one smoker; (B) households that are not eligible or eligible for healthy start vouchers. Averages +/-SE. *** p<0.001; ** p<0.01

## DISCUSSION

The public health and economic burden of CO exposure remains unknown: CO-related health issues are likely underdiagnosed due to the nonspecific symptoms of exposure, and CO is not regularly assessed in the home (or, indeed, in the environment) as current safety guidelines are generally assumed to be met. It is furthermore not certain what are the safe levels for pregnant women and the vulnerable foetus. We sought to quantify CO levels in the homes of pregnant women, identify effective indicators of home-based CO exposure, and assess whether exhaled CO values could predict home-based CO exposure.

### Exposure levels

We detected CO in the households of the majority of pregnant women enrolled in our study, with a third and a sixth of households experiencing CO levels above 4ppm and 8ppm, respectively. The pattern of exposure varied greatly between households, most likely due to the underlying source of CO emission. While almost half of households had exposures lasting 10 minutes or more, an equally typical pattern was repeated exposures, with significant peaks followed by periods of little or no CO. Although CO exposure was widespread, most of the households studied were compliant with current guidelines.

Studies mapping CO exposure in different subsets of the UK population have shown similar results, indicating that CO is present in a number of households, but not necessarily at levels exceeding guidelines. In a large study of >800 households, there were no incidences of exceeding an earlier WHO guideline of 8.6ppm, however maximum levels reported indicated peaks of 3.88ppm (Raw et al., 2004), considerably lower than the average maximum level detected here (10.4+/-33.5ppm). However, a study in East London of 270 households from showed that 18% exhibited CO levels exceeding the then WHO guidelines of 8.6ppm (Croxford et al., 2005). Similarly, a study in 20 non-smoking households and 44 smoking households found that although smoking households had a higher mean (0.1-21ppm) and maximum exposure level (1.9-53.6ppm) than non-smoking households (mean: 0.1-1ppm; maximum: 4-22ppm), neither group exhibited means in excess of 10ppm over an 8-hr period (Henderson et al., 2006). In this case the sample was selected on the basis of age, which might limit potential comparisons with our study. Nevertheless, it is interesting to observe that maximum exposures were on par with those identified here.

### Source of CO

Likely sources of CO in the home are cigarette smoking and gas appliances, the presence of which might more than double the CO levels in the home (Green et al., 1999; World Health Organization, 1999). We found that CO levels fluctuated across the 24-hr period, both mean and max values varying with a polynomial (cubic) fit. Levels were typically higher at night, with max values also rising around 4pm-6pm. We cannot formally test the cause of these fluctuations, as participants were not asked to record their day-to-day activities, but smoking is unlikely to be the main driver for these fluctuations given their elevation at night. A more likely scenario is that CO levels are linked to gas heaters (heating turned on as the temperature drops in the evening, remaining on until the morning; heating turned off during the day as people may be at work) and gas cooking (around 17:00-18:00), both appliances known to emit CO. While we could not assess correlation between gas heaters and the other variables in this study, due to too few households not being heated by gas (reflecting our inclusion criteria), it is important to note that this does not rule out an impact of gas heating on our measured CO levels. Importantly, we did see a significant correlation between using gas for cooking and all measured CO values in the home in the present study, which greatly supports the above interpretation and supports a similar finding from an earlier study (Raw et al., 2004). It is also important to note that CO concentration in the immediate vicinity of the cooker will likely be higher than what might that detected by the loggers, and traditionally, women are more likely to be involved in food preparation, hence more at risk of exposure.

### Risk factors

We observed correlations between exhaled CO levels at visit 1 and several factors, including maximal CO exposure (as measured by the CO data loggers), voucher eligibility and number of smokers in the property, type of occupancy and ethnicity. Exhaled CO levels remain a simple way of assessing CO exposure in pregnancy, and linking this to risk factors may thus help to identify vulnerable sections of the population. Indeed, both voucher eligibility and number of smokers were significant predictors for CO levels measured in the exhaled breath. Taken together, this supports other studies suggesting increased risk of CO exposure for deprived areas. For example, in a study on low-income households, fuel poverty was shown to be a risk factor for elevated CO levels in the home (NEA, 2017).

A key aim of our study was to ask whether breath CO measurements could indicate environmental exposures other than smoking. Notably, breath measurements did not emerge as a correlate of CO indices collected by the CO alarms or data loggers, nor did household CO levels predict CO breath levels. As household CO levels in the study were generally well within recommended limits, it is possible that CO breath tests may only be suitable for detecting exposures exceeding current safety guidelines. CO exposures outside the home (which we did not record) may also have introduced further variation into our measurements, confounding our analysis. Indeed, there is no straightforward translation between exposures and CO concentration in the body, although there have been several attempts at generating an algorithm for CO uptake (Tikuisis, 1996). Despite this result, it is important to note that elevated CO in a breath test cannot be presumed to be caused by exposure to tobacco smoke.

CO exposure is difficult to diagnose without a source of the gas being identified, as the gas typically cause non-specific symptoms such as dizziness, nausea and headache. Depending on the exposure level, the duration of the exposure, and the specific physical condition of the person exposed, CO could cause health implications not only in the short term, but also persisting over time (Ashcroft et al., 2019; Smollin and Olson, 2008; Knobeloch and Jackson, 1999). Indeed, evidence is starting to emerge that even very low exposures may impact on health (Bendell et al., 2020; Wilson & Herigstad, 2024), particularly in vulnerable populations such as the developing foetus (Matias et al., 2024). Notably, exposure to CO at air pollution levels, which are comparable to levels measured in the present study, has been linked to higher incidences of dementia (Chang et al., 2014), stroke (Hedblad et al., 2005; Maheswaran et al., 2005) and heart failure (Shah et al., 2013), to altered brain vascular function (Bendell et al., 2020) and remodelling of the developing heart (Matias et al., 2024). It is thus imperative to improve our understanding of risk factors and prevalence of CO exposure.

### CO in pregnancy

The foetus is particularly vulnerable to CO exposure. CO readily crosses the placenta, and as a result of several factors including the higher affinity of foetal Hb for CO, the foetus will typically experience higher maximal values of COHb (Lopez et al., 2015) that persist for longer periods compared to the mother (Aubard & Magne, 2000). E.g. if maternal exhaled CO level is 13ppm, as can be seen in a typical smoker, this would on average correspond to a maternal % COHb of 2.08% but a foetal %FCOHb of 4.07% (Lopez et al., 2015).

Case reports of maternal CO poisoning are fortunately rare, but highlight a range of serious adverse foetal outcomes, including cerebral palsy (Alehan et al., 2007), hypoxic ischemic encephalopathy (Tuoni et al., 2023; Yildiz et al., 2010) and cardiomegaly (Aubard & Magne, 2000) as well as death (Yildiz et al., 2010). Maternal smoking is known to carry increased risk of miscarriage, stillbirth, prematurity, foetal growth restriction and low birth weight (Cnattingius, 2004; Herrmann et al., 2008; Pineles et al., 2014). There is also evidence that maternal smoking may cause neurocognitive and behavioural effects in the child (Batstra et al., 2003; Herrmann et al., 2008). As cigarette smoke contains a cocktail of toxins, it remains unclear how much of the adverse effects of maternal smoking can be attributed to CO alone.

However, work in rats has shown that exposure to CO at levels associated with smoking causes reduced foetal birth weight, but nicotine or aldehydes (also present in tobacco smoke) do not have a similar effect (Carmines & Rajendran, 2008). Such work has also confirmed the neurological sequelae of CO exposure: maternal exposure to 75-150ppm CO in rats irreversibly impairs learning ability and memory in the offspring (De Salvia et al., 1995; Giustino et al., 1999). Furthermore, maternal exposure to second-hand smoke has been associated with impaired motor ability (Hernández-Martínez et al., 2012) and neurodevelopmental delay (Lee et al., 2011). Lee et al. furthermore observed a decrease in mental developmental index score (which incorporates attentiveness and response to stimulation) in children (6 months) whose mothers had been exposed to second-hand smoke during pregnancy, even after adjusting for covariates such as residential area, maternal age and education, income and birth weight (Lee et al., 2011).

There is some evidence that maternal exposure to CO at even lower doses than those associated with smoking carries an increased risk of several adverse developmental outcomes. For instance, maternal chronic exposure to CO through woodsmoke at levels of 12.5ppm and below has been linked to lower neuropsychological test scores, visuo-spatial integration and motor performance in children aged 6-7 years (Dix-Cooper et al., 2012). Epidemiological associations between birth weight reduction and environmental CO increases of 1.4ppm (Salam et al., 2005), or levels of 5.5ppm and above (Ritz & Yu, 1999), have been reported, to mention but two. Multiple studies have reported higher odds of low foetal size or birth weight with low single figure increases in CO concentrations (for example Thompson et al., 2011; Yucra et al., 2014). Animal studies have not typically been performed at such low doses, but in a chick model of development, levels as low as 3.5ppm over the first 9 days of gestation has been shown to enlarge the heart walls and ventricular septum, and the study further demonstrated this this occurred in a dose-dependent manner (Matias et al., 2024). Overall, these data are in line with the assumption that both the heart and brain are particularly vulnerable to CO. Indeed, only ∼30% of congenital heart defects are currently explicable through genetics, with much of the remaining variation thought to be due to environmental factors (Kalisch-Smith et al., 2021). As cardiac defects are the most prevalent type of birth defect, even a slight increase in risk could translate to a high global burden of morbidity. It thus remains unclear at what concentration CO exposure no longer presents a health risk.

### Limitations

Recruitment was conducted from maternity services at University Hospital Coventry & Warwickshire, Royal United Hospital Bath, Queen Elizabeth Hospital Gateshead, Mid Yorkshire NHS Trust (Pinderfields and Pontefract Hospitals) and Northumbria Hospital. While no selection for socioeconomic factors was conducted during recruitment, these hospitals do serve deprived populations, which may skew the findings towards higher CO exposures. Future studies might want to include a greater variety of study sites to assess differences in sociodemographic factors with greater accuracy.

## Conclusion

This study shows that the CO exposure levels in a subset of UK households is higher than current guidelines, and that this can be linked to the use of gas in cooking. The highest of the CO levels observed are on par with doses that have produced adverse foetal outcomes both in epidemiological studies in humans and in model organisms in the laboratory. This study also shows that exhaled CO levels in pregnant women can be predicted by the number of smokers in the household and eligibility for the Healthy Start scheme, but that the latter association may potentially be modified (possibly by CO awareness) as it was no longer present at the second visit. In conclusion, we argue that CO exposure remains a healthcare challenge in the pregnant population, which could have tangible public health consequences, and that further study as well as awareness campaigns are warranted, particularly in at-risk populations, to address this issue.

## Data Availability

All data produced in the present study are available upon reasonable request to the authors

## Acknowledgements

This study was funded by the CO Research Trust and the Gas Distribution Networks that covered the study sites.

## Notes

### Competing Interest Statement

The authors have declared no competing interest.

### Author Declarations

The Integrated Research Application System (IRAS) gave ethical approval for this work (IRAS ID 301261; East Midlands - Nottingham 1 Research Ethics Committee).

## References

Alehan, F., Erol, I., & Onay, Ö. S. (2007). Cerebral palsy due to nonlethal maternal carbon monoxide intoxication. Birth Defects Research Part A - Clinical and Molecular Teratology, 79(8), 614–616. 10.1002/bdra.20379

Aubard, Y., & Magne, I. (2000). Carbon Monoxide Poisoning in Pregnancy. American Journal of Obstetrics and Gynecology, 107, 833–838.

Batstra, L., Hadders-Algra, M., & Neeleman, J. (2003). Effect of antenatal exposure to maternal smoking on behavioural problems and academic achievement in childhood: Prospective evidence from a Dutch birth cohort. Early Human Development, 75(1–2), 21–33. 10.1016/j.earlhumdev.2003.09.001

Bendell, C., Moosavi, S. H., & Herigstad, M. (2020). Low-level carbon monoxide exposure affects BOLD fMRI response. Journal of Cerebral Blood Flow and Metabolism, 40(11), 2215–2224. 10.1177/0271678X19887358

Cagiano, R., Ancona, D., Cassano, T., Tattoli, M., Trabace, L., & Cuomo, V. (1998). Effects of prenatal exposure to low concentrations of carbon monoxide on sexual behaviour and mesolimbic dopaminergic function in rat offspring. British Journal of Pharmacology, 125(4), 909–915. 10.1038/sj.bjp.0702143

Carmines, E. L., & Rajendran, N. (2008). Evidence for carbon monoxide as the major factor contributing to lower fetal weights in rats exposed to cigarette smoke. Toxicological Sciences, 102(2), 383–391. 10.1093/toxsci/kfn009

Chang, K. H., Chang, M. Y., Muo, C. H., Wu, T. N., Chen, C. Y., & Kao, C. H. (2014). Increased risk of dementia in patients exposed to nitrogen dioxide and carbon monoxide: a population-based retrospective cohort study. PLoS One, 9(8), e103078. 10.1371/journal.pone.0103078

Cnattingius, S. (2004). The epidemiology of smoking during pregnancy: Smoking prevalence, maternal characteristics, and pregnancy outcomes. Nicotine and Tobacco Research, 6(SUPPL. 2). 10.1080/14622200410001669187

Croxford, B., Hutchinson, E., Leonardi, G. S., Mckenna, L., Nicholson, L., Volans, G., & Wilkinson, P. (2005). Real time carbon monoxide measurements from 270 UK homes. In J. J. (Ed.), Proceedings of Indoor Environmental Quality (IEQ) – Problems, Research, and Solutions. USA:Air and Waste Management Association (A&WMA) and US EPA’s Office of Research and Development.

Dadvand, P., Rankin, J., Rushton, S., & Pless-Mulloli, T. (2011). Association between maternal exposure to ambient air pollution and congenital heart disease: A register-based spatiotemporal analysis. Am J Epidemiol, 173(2), 171–182. 10.1093/aje/kwq342

De Salvia, M. A., Cagiano, R., Carratù, M. R., Di Giovanni, V., Trabace, L., & Cuomo, V. (1995). Irreversible impairment of active avoidance behavior in rats prenatally exposed to mild concentrations of carbon monoxide. Psychopharmacology, 122(1), 66–71. 10.1007/BF02246443

Di Giovanni, V., Cagiano, R., De Salvia, M., Giustino, A., Lacomba, C., Renna, G., & Cuomo, V. (1993). Neurobehavioral changes produced in rats by prenatal exposure to carbon monoxide V i n c e n. Brain Research, 616, 126–131.

Dix-Cooper, L., Eskenazi, B., Romero, C., Balmes, J., & Smith, K. R. (2012). Neurodevelopmental performance among school age children in rural Guatemala is associated with prenatal and postnatal exposure to carbon monoxide, a marker for exposure to woodsmoke. NeuroToxicology, 33(2), 246–254. 10.1016/j.neuro.2011.09.004

Giustino, A., Cagiano, R., Carratù, M. R., Cassano, T., Tattoli, M., & Cuomo, V. (1999). Prenatal exposure to low concentrations of carbon monoxide alters habituation and non-spatial working memory in rat offspring. Brain Research, 844(1–2), 201–205. 10.1016/S0006-8993(99)01832-6

Green, E., Short, S., Shuker, L. K., & Harrison, P. T. C. (1999). Carbon monoxide exposure in the home environment and the evaluation of risks to health - A UK perspective. Indoor and Built Environment, 8(3), 168–175. 10.1159/000024632

Hedblad, B., Ogren, M., Engstrom, G., Wollmer, P., & Janzon, L. (2005). Heterogeneity of cardiovascular risk among smokers is related to degree of carbon monoxide exposure. Atherosclerosis, 179(1), 177–183. 10.1016/j.atherosclerosis.2004.10.005

Henderson, K. A., Parry, S., & Matthews, I. A. N. P. (2006). Real-time measurement of short-term peaks in environmental CO concentrations in the homes of the elderly in South Wales. 525–530. 10.1038/sj.jes.7500491

Hernández-Martínez, C., Arija Val, V., Escribano Subías, J., & Canals Sans, J. (2012). A longitudinal study on the effects of maternal smoking and secondhand smoke exposure during pregnancy on neonatal neurobehavior. Early Human Development, 88(6), 403–408. 10.1016/j.earlhumdev.2011.10.004

Herrmann, M., King, K., & Weitzman, M. (2008). Prenatal tobacco smoke and postnatal secondhand smoke exposure and child neurodevelopment. Current Opinion in Pediatrics, 20(2), 184–190. 10.1097/MOP.0b013e3282f56165

Kalisch-Smith, J. I., Ved, N., Szumska, D., Munro, J., Troup, M., Harris, S. E., Rodriguez-Caro, H., Jacquemot, A., Miller, J. J., Stuart, E. M., Wolna, M., Hardman, E., Prin, F., Lana-Elola, E., Aoidi, R., Fisher, E. M. C., Tybulewicz, V. L. J., Mohun, T. J., Lakhal-Littleton, S., … Sparrow, D. B. (2021). Maternal iron deficiency perturbs embryonic cardiovascular development in mice. Nature Communications, 12(1), 1–17. 10.1038/s41467-021-23660-5

Kokkarinen, N., Shaw, A., Cullen, J., Pedrola, M. O., Mason, A., & Al-Shamma’a, A. (2014). Investigation of audible carbon monoxide alarm ownership□: Case study. Smart and Sustainable Built Environment, 3(1), 72–86. 10.1108/SASBE-07-2013-0041

Lee, B. E., Hong, Y. C., Park, H., Ha, M., Hyeong Kim, J., Chang, N., Roh, Y. M., Kim, B. N., Kim, Y., Oh, S. young, Ju Kim, Y., & Ha, E. H. (2011). Secondhand smoke exposure during pregnancy and infantile neurodevelopment. Environmental Research, 111(4), 539–544. 10.1016/j.envres.2011.02.014

Longo, L. D. (1977). The biological effects of carbon monoxide on the pregnant woman, fetus, and newborn infant. Am J Obstet Gynecol, 129(1), 69–103. 10.1016/0002-9378(77)90824-9

Lopez, A. S., Waddington, A., Hopman, W. M., & Jamieson, M. A. (2015). The Collection and Analysis of Carbon Monoxide Levels as an Indirect Measure of Smoke Exposure in Pregnant Adolescents at a Multidisciplinary Teen Obstetrics Clinic. Journal of Pediatric and Adolescent Gynecology, 28(6), 538–542. 10.1016/j.jpag.2015.04.007

Maheswaran, R., Haining, R. P., Brindley, P., Law, J., Pearson, T., Fryers, P. R., Wise, S., & Campbell, M. J. (2005). Outdoor air pollution and stroke in Sheffield, United Kingdom: a small-area level geographical study. Stroke, 36(2), 239–243. 10.1161/01.STR.0000151363.71221.12

Matias, F. R., Groves, I., Durrans, J., & Herigstad, M. (2024). Carbon monoxide affects early cardiac development in an avian model. Birth Defects Research, 116(3), 1–11. 10.1002/bdr2.2330

NEA. (2017). Understanding carbon monoxide risk in households vulnerable to fuel poverty. National Energy Action.

NICE. (2010). Smoking: Stopping in pregnancy and after childbirth. NICE Guidelines PH26. June 2010. https://www.nice.org.uk/guidance/PH26

Penney, D., Benignus, V., Kephalopoulos, S., Kotzias, D., Kleinman, M., & Verrier, A. (2010). Carbon Monoxide. In WHO Guidelines for Indoor Air Quality: Selected Pollutants (pp. 55–102).

Pineles, B. L., Park, E., & Samet, J. M. (2014). Systematic review and meta-analysis of miscarriage and maternal exposure to tobacco smoke during pregnancy. American Journal of Epidemiology, 179(7), 807–823. 10.1093/aje/kwt334

Raub, J. A., & Benignus, V. A. (2002). Carbon monoxide and the nervous system. Neuroscience and Biobehavioral Reviews, 26(8), 925–940. 10.1016/S0149-7634(03)00002-2

Raw, G. J., Coward, S. K. D., Brown, V. M., & Crump, D. R. (2004). Exposure to air pollutants in English homes. 10.1038/sj.jea.7500363

Ritz, B., & Yu, F. (1999). The Effect of Ambient Carbon Monoxide on Low Birth Weight among Children Born in Southern California between 1989 and 1993. Environmental Health Perspectives, 107(1), 17–25.

Ritz, B., Yu, F., Fruin, S., Chapa, G., Shaw, G. M., & Harris, J. A. (2002). Ambient air pollution and risk of birth defects in Southern California. Am J Epidemiol, 155(1), 17–25. 10.1093/aje/155.1.17

Salam, M. T., Millstein, J., Li, Y. F., Lurmann, F. W., Margolis, H. G., & Gilliland, F. D. (2005). Birth outcomes and prenatal exposure to ozone, carbon monoxide, and particulate matter: Results from the Children’s Health Study. Environmental Health Perspectives, 113(11), 1638–1644. 10.1289/ehp.8111

Shah, A. S., Langrish, J. P., Nair, H., McAllister, D. A., Hunter, A. L., Donaldson, K., Newby, D. E., & Mills, N. L. (2013). Global association of air pollution and heart failure: a systematic review and meta-analysis. Lancet, 382(9897), 1039–1048. 10.1016/S0140-6736(13)60898-3

Smollin, C., & Olson, K. (2008). Carbon monoxide poisoning (acute). BMJ Clin Evid, 7, 1–12.

Thom, S. R., Taber, R. L., Mendiguren II, Clark, J. M., Hardy, K. R., & Fisher, A. B. (1995). Delayed neuropsychologic sequelae after carbon monoxide poisoning: prevention by treatment with hyperbaric oxygen. Ann Emerg Med, 25(4), 474–480. 10.1016/s0196-0644(95)70261-x

Thompson, L. M., Bruce, N., Eskenazi, B., Diaz, A., Pope, D., & Smith, K. R. (2011). Impact of reduced maternal exposures to wood smoke from an introduced chimney stove on newborn birth weight in rural Guatemala. Environmental Health Perspectives, 119(10), 1489–1494. 10.1289/ehp.1002928

Tikuisis, P. (1996). Modeling the Uptake and Elimination of Carbon Monoxide. In D. G. Penney (Ed.), Carbon Monoxide. Taylor & Francis.

Townsend, C. L., & Maynard, R. L. (2002). Effects on health of prolonged exposure to low concentrations of carbon monoxide. Occup Environ Med, 59(10), 708–711. 10.1136/oem.59.10.708

Trentini, J. F., O’Neill, J. T., Poluch, S., & Juliano, S. L. (2016). Prenatal carbon monoxide impairs migration of interneurons into the cerebral cortex. NeuroToxicology, 53, 31–44. 10.1016/j.neuro.2015.11.002

Tuoni, C., Nuzzi, G., Scaramuzzo, R. T., Fiori, S., & Filippi, L. (2023). Neonatal hypoxic-ischemic encephalopathy after acute carbon monoxide intoxication during pregnancy. A case report and brief review of the literature. Frontiers in Pediatrics, 11(November), 1–5. 10.3389/fped.2023.1264855

Venditti, C. C., Casselman, R., Murphy, M. S. Q., Adamson, S. L., Sled, J. G., & Smith, G. N. (2013). Chronic carbon monoxide inhalation during pregnancy augments uterine artery blood flow and uteroplacental vascular growth in mice. American Journal of Physiology-Regulatory Integrative and Comparative Physiology, 305(8). 10.1152/ajpregu.00204.2013

Wilson, L. A., & Herigstad, M. (2024). Impact of carbon monoxide on neural activation during a reaction time task. BioRxiv. 10.1101/2023.01.17.524443

World Health Organization. (1999). Air quality guidelines for Europe. Chapter 5.5, carbon monoxide. https://www.euro.who.int/data/assets/pdf_file/0020/123059/AQG2ndEd_5_5carbonmonoxide.PDF

Yildiz, H., Aldemir, E., Altuncu, E., Celik, M., & Kavuncuoglu, S. (2010). A rare cause of perinatal asphyxia: Maternal carbon monoxide poisoning. Archives of Gynecology and Obstetrics, 281(2), 251–254. 10.1007/s00404-009-1139-4

Yucra, S., Tapia, V., Steenland, K., Naeher, L. P., & Gonzales, G. F. (2014). Maternal exposure to biomass smoke and carbon monoxide in relation to adverse pregnancy outcome in two high altitude cities of Peru. Environmental Research, 130, 29–33. 10.1016/j.envres.2014.01.008

